## Supplementary Figures 1-6 for "Examining the association between fetal *HLA-C*, maternal *KIR* haplotypes and birth weight"

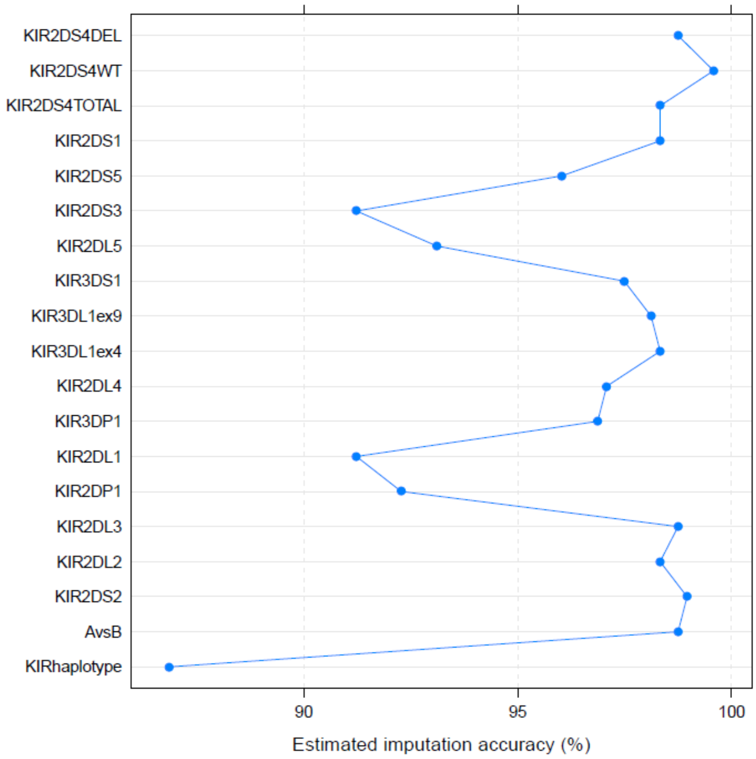


**a)**

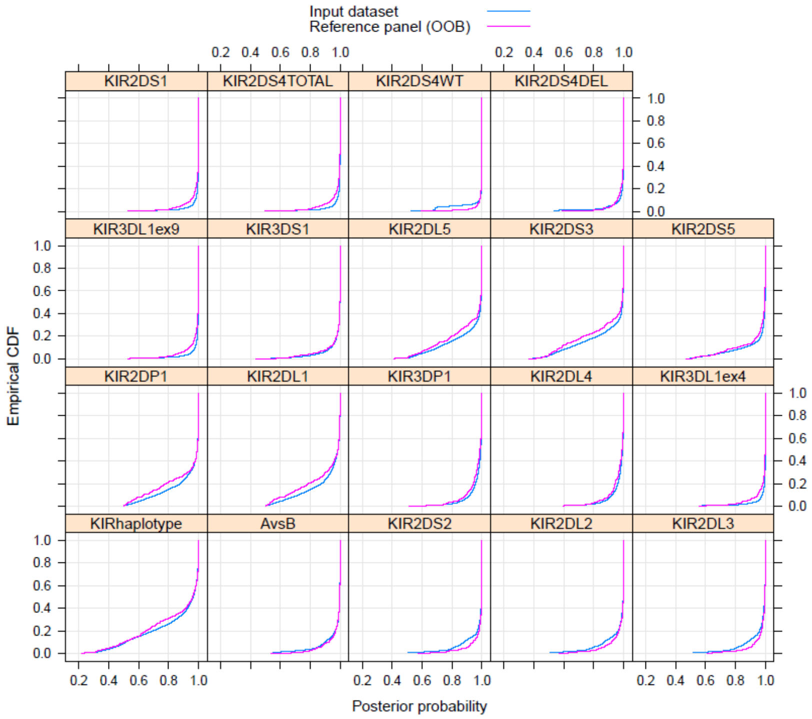


**b)**

**Supplementary Figure S1.** KIR imputation plots for UK Biobank (UKB) mother-offspring pairs (n=8,498 at imputation stage) showing **a)** the estimated imputation accuracy achieved at each KIR locus and **b)** the distribution of the posterior probabilities of the most likely alleles for each KIR locus where the distribution of the input dataset is shown in blue against the reference panel distribution shown in red.

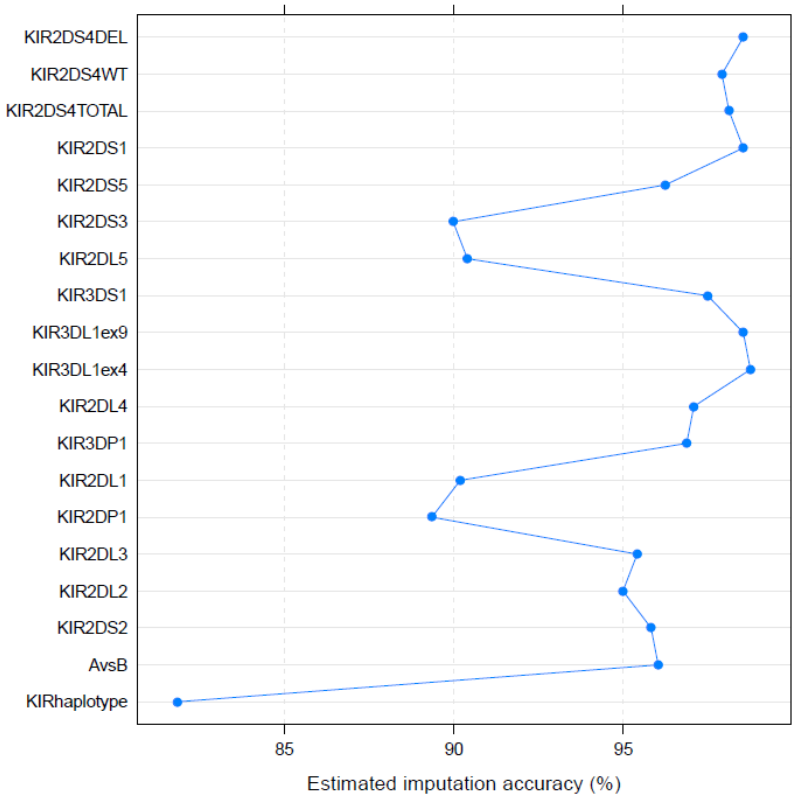


**a)**

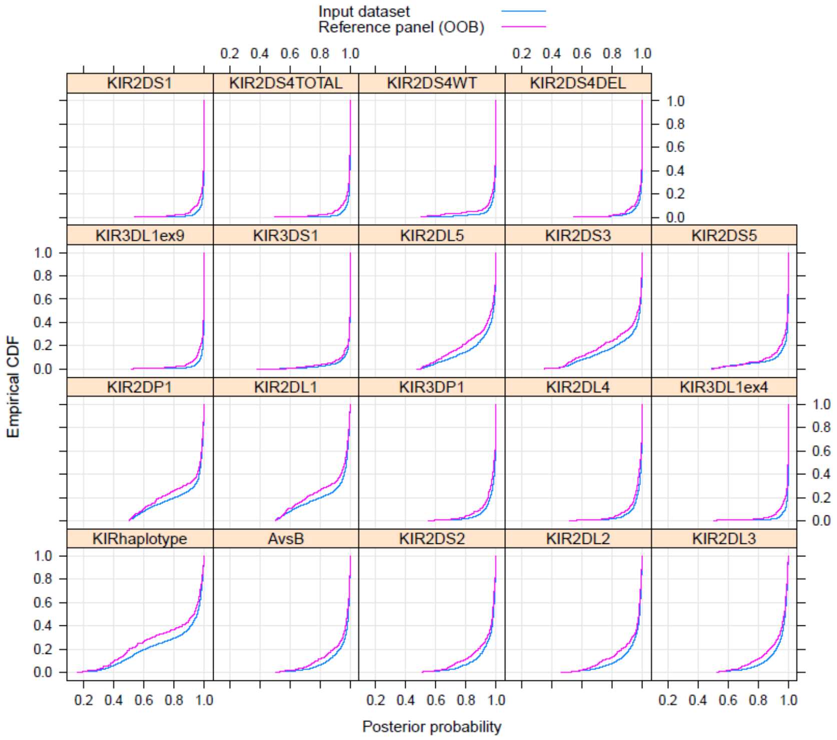


**b)**

**Supplementary Figure S2.** KIR imputation plots for the Exeter Family Study of Childhood Health (EFSOCH) mother-offspring pairs (n=2,664 at imputation stage) showing **a)** the estimated imputation accuracy achieved at each KIR locus and **b)** the distribution of the posterior probabilities of the most likely alleles for each KIR locus where the distribution of the input dataset is shown in blue against the reference panel distribution shown in red.

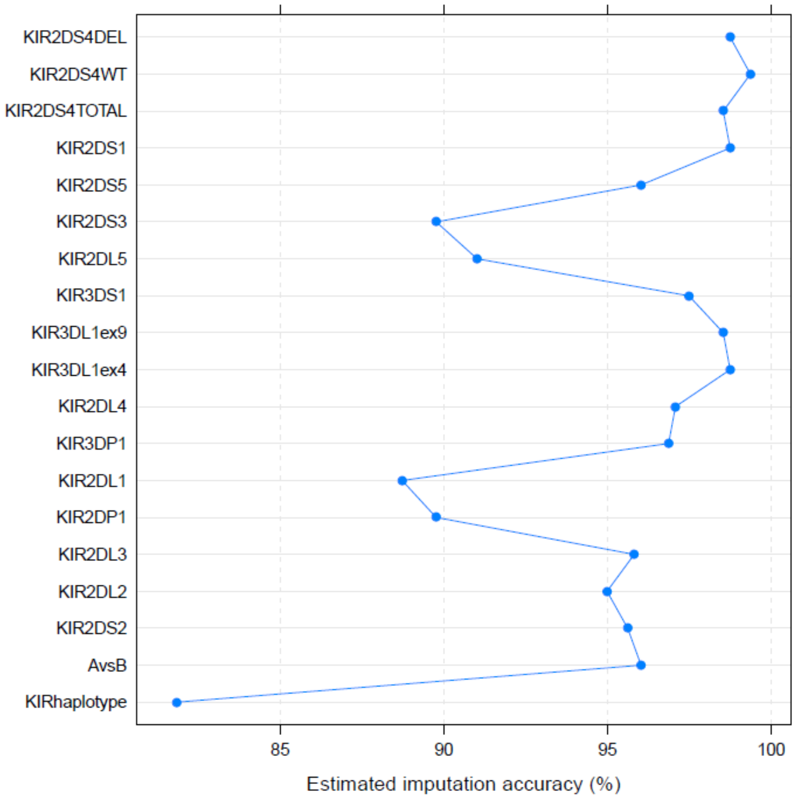


**a)**

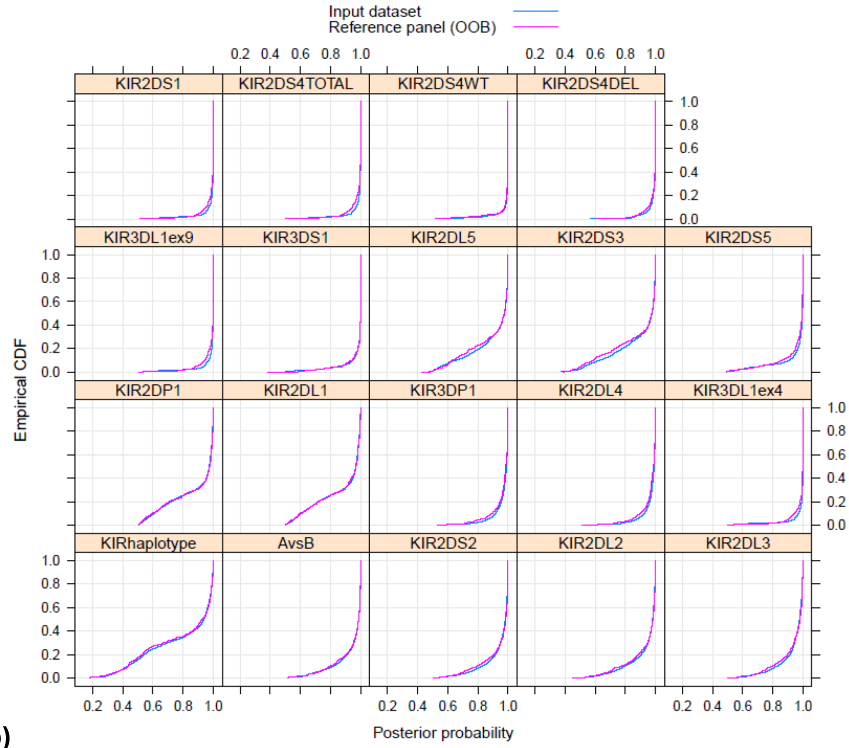


**b)**

**Supplementary Figure S3.** KIR imputation plots for the Hyperglycemia and Adverse Pregnancy Outcome (HAPO) study mother-offspring pairs (n=1,734 at imputation stage) showing **a)** the estimated imputation accuracy achieved at each KIR locus and **b)** the distribution of the posterior probabilities of the most likely alleles for each KIR locus where the distribution of the input dataset is shown in blue against the reference panel distribution shown in red.

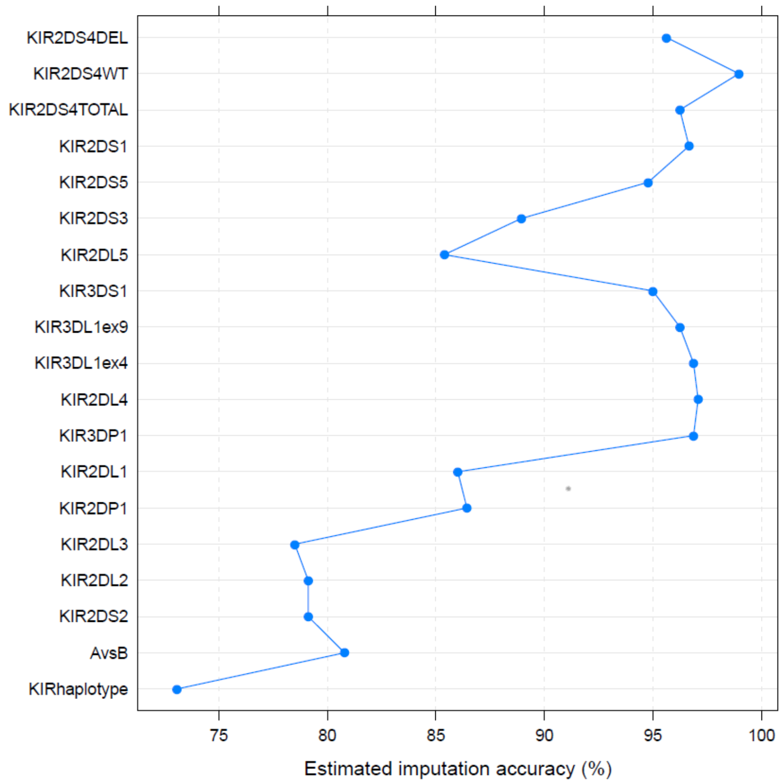


**a)**

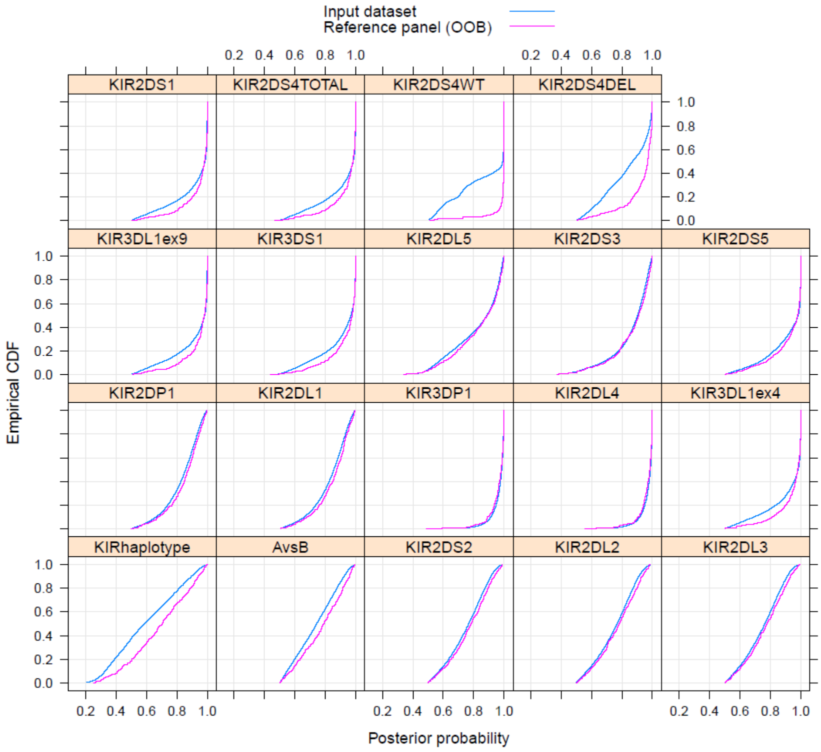


**b)**

**Supplementary Figure S4.** KIR imputation plots for the Avon Longitudinal Study of Parents and Children (ALSPAC) mother-offspring pairs (n=17,827 at imputation stage) showing **a)** the estimated imputation accuracy achieved at each KIR locus and **b)** the distribution of the posterior probabilities of the most likely alleles for each KIR locus where the distribution of the input dataset is shown in blue against the reference panel distribution shown in red.

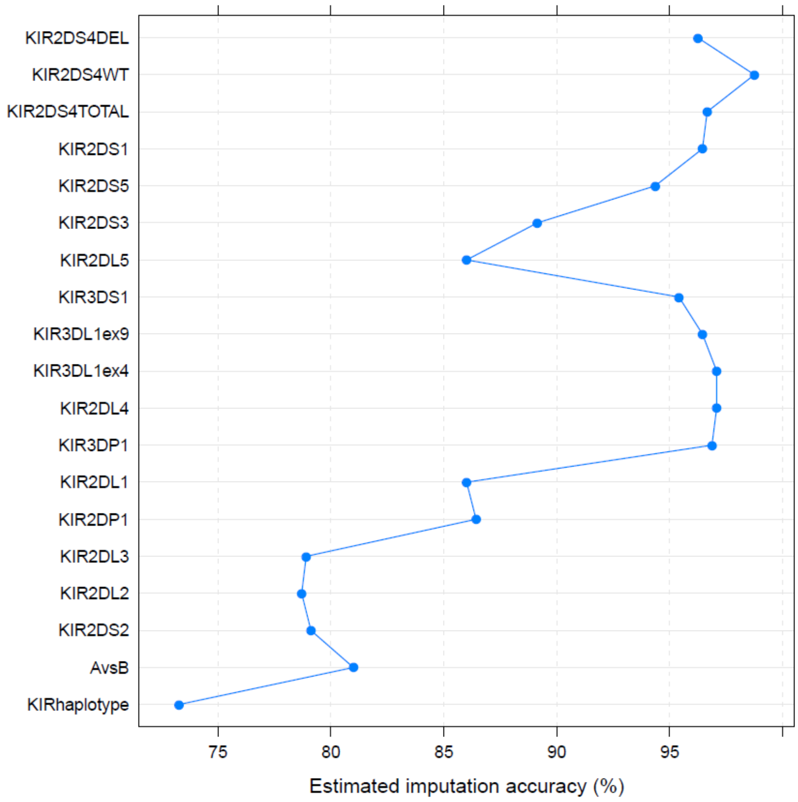


**a)**

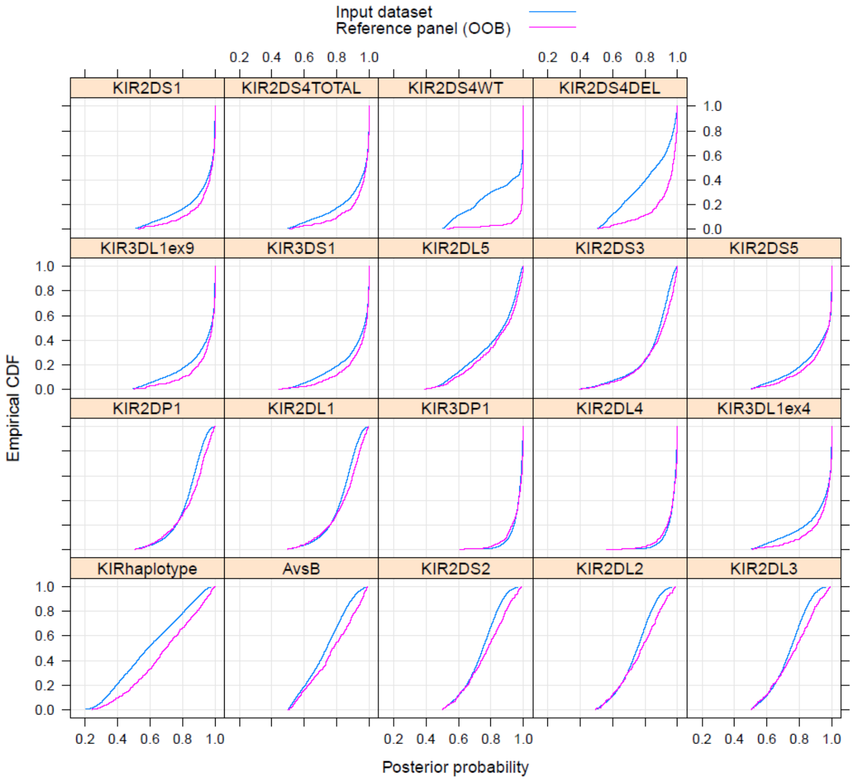


**b)**

**Supplementary Figure S5.** KIR imputation plots for the Born in Bradford (BiB) Core Exome mother-offspring pairs (n=5,709 at imputation stage) showing **a)** the estimated imputation accuracy achieved at each KIR locus and **b)** the distribution of the posterior probabilities of the most likely alleles for each KIR locus where the distribution of the input dataset is shown in blue against the reference panel distribution shown in red.

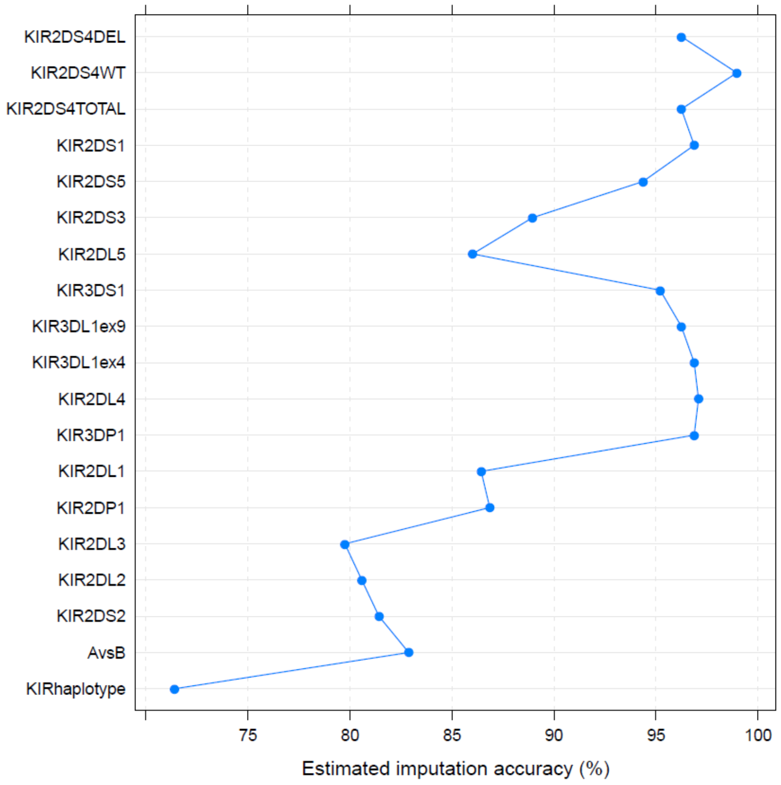


a)

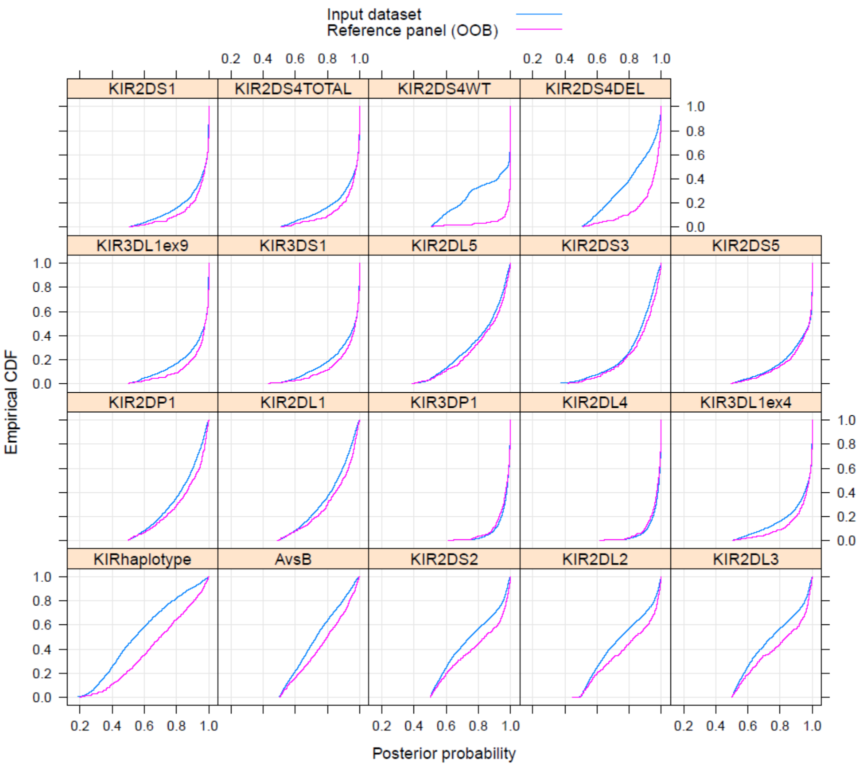


b)

**Supplementary Figure S6.** KIR imputation plots for the Born in Bradford (BiB) Global Screening Array mother-offspring pairs (n=1,901 at imputation stage) showing **a)** the estimated imputation accuracy achieved at each KIR locus and **b)** the distribution of the posterior probabilities of the most likely alleles for each KIR locus where the distribution of the input dataset is shown in blue against the reference panel distribution shown in red.
